# Leveraging Pharmacy Education through a Train-the-Trainer Model to Enhance Breast Cancer Literacy in Rural Communities

**DOI:** 10.1101/2025.03.25.25324513

**Authors:** Christopher L Farrell, Melanie R Ginzburg, Morgan B Enlow, Missouri M Jenkins, Alexus N Hames, Cayla R Adams, Marlana A Roberts, Hillary E Stamps, Natalie A Paxton, Courtney N Addison, Austin Y Shull

## Abstract

Rural versus urban communities experience disproportionate challenges in breast cancer outcomes, with higher breast cancer mortality and later stage disease presentation, despite similar diagnosis rates. These disparities are driven by structural barriers, including rural hospital closures, transportation difficulties, and limited access to oncology specialists. This study evaluated a train-the-trainer program designed to equip PharmD students located at a pharmacy school in a rural county in South Carolina with breast cancer education training, leveraging the pharmacists’ position as accessible healthcare professionals in rural communities. Training focused on breast cancer risk factors, prevention, screening, genetics, staging, and treatment options. Effectiveness was measured through pre- and post-workshop confidence surveys and knowledge assessments. Results showed significant improvement in student confidence across educational domains, with average scores increasing from 6.30 to 8.59 (p<0.0001). Understanding of screening guidelines (mean difference: 4.30; p-value: <.0001) and target therapy options showed the greatest improvement (mean difference: 3.65; p-value: <.0001), while knowledge of BRCA gene inheritance showed the smallest change (mean difference: 0.369; p-value: ns), suggesting some pre-existing awareness but limited understanding of its clinical applications. Overall, this pilot program demonstrates how pharmacy education can address healthcare disparities in rural communities. By preparing pharmacists to deliver accurate breast cancer education and to increase rural patient agency, this model creates a sustainable approach to improving health literacy in medically underserved areas. Future research could further expand this model to include diverse healthcare professionals and incorporate long-term impact assessments in community settings.

## INTRODUCTION

The importance of access to medical literacy and education has been a strong focus in recent years, particularly in rural and lower socioeconomic communities, where a substantial portion of medically underserved populations reside^1^. These communities face unique challenges, such as a scarcity of resources, that limit access to quality healthcare and hinder their ability to make informed health-related decisions. Among these unique health concerns, breast cancer is as a particularly pressing issue, since it is the second leading cause of cancer death among women in the United States, and its mortality rate is disproportionately higher among Black and Hispanic women who make up a significantly growing portion of the rural community^2,3^. While breast cancer diagnoses are not significantly different between urban and rural areas, recent studies reveal that late-stage breast cancer presentation positively correlated with rurality, as did higher breast cancer mortality in women from rural areas in the Southern United States^4–6^. Such structural barriers driving these disproportional outcomes include limited access to oncology specialists and treatment centers, the recent increase of hospital closures in rural communities, and transportation to appropriate medical facilities that are concentrated in urban areas. These geographically associated barriers are large contributors to rural residents receiving less frequent cancer screenings and more late-stage cancer diagnoses^7^.

These disparities underscores the importance of implementing efforts to bridge the gap in medical literacy and education access for rural patients to increase their advocacy and improve their personal outcomes in chronic conditions like breast cancer. This enhanced medical literacy can contribute to agency that then enables better management of chronic conditions and encouraging optimal use of healthcare services like valuing regular check-ups, preventive screenings, and early intervention. Consequently, by providing accurate and comprehensible health information, healthcare providers can in turn work toward minimizing disparities in healthcare access, especially based on knowledge of healthcare programs that are designed to address these issues within rural communities, thus actively promote better health equity in underserved communities^8–12^. Therefore, this dual partnership between healthcare professional and patient enhances meaningful communication between individuals who understand medical terminology, treatment options, and potential risks and fosters a stronger patient-provider relationship that leads to better treatment adherence and outcomes.

However, a critical bottleneck in this pursuit is the availability of a suitable cohort equipped with a foundational medical education to seamlessly facilitate these medical literacy, education, and advocacy approaches within rural communities in a way that is consistent and fosters trust within a community^13^. This limitation highlights the necessity for innovative approaches to equip individuals with the requisite skills and knowledge to serve as conduits of accurate information and support^14^. In response to this challenge, we have developed and evaluated a “train-the-trainer” program, a strategy that entails imparting health professional graduate students, particularly those pursuing PharmD degrees, with the necessary training and resources^15^. While the training provided could be utilized in many health professional programs, we specifically focused on the PharmD program based on the understanding that pharmacists are one of most accessible healthcare providers, especially in rural communities^16–18^. In fact, statistics show that an average patient visits a pharmacy nearly 2-3 times more often than a primary care provider, therefore allowing for more opportunity to build patient trust and connectivity for medical education, disease management, and over health advocacy opportunities^19^. Given this preparation to provide effective breast cancer education to their rural patients and communities, these healthcare professional PharmD students through our breast cancer education “train-the-trainer” program can be equipped to effectively implement breast cancer education outreach programs and thereby foster a more informed and empowered patient basis that can address the rural disparities in breast cancer outcomes.

## METHODS

The protocol for this study was approved by Presbyterian College’s Institutional Review Board. For this training-based study, two breast cancer education workshops were conducted between 2018 and 2019 for the purposes of preparing second and third year PharmD students from the Presbyterian College School of Pharmacy to educate their respective communities on breast cancer risk, prevention, and treatment. Presbyterian College School of Pharmacy serves as an ideal educational site for this training, since it resides in Laurens County, South Carolina (SC). Laurens County and its neighboring Greenwood and Union Counties, also in SC, are defined as rural counties by the Health Services Resources and Services Administration [HRSA] (hrsa.gov). Their poverty rates (16.7 percent [Laurens County, SC], 16.9 percent [Greenwood County, SC], and 23.4 percent [Union County, SC]), are all considerably higher than that for the state (13.9 percent) and the nation (11.1 percent) (census.gov).

Professors with extensive PhD training in oncology designed a PowerPoint-style presentation (Supplementary File S1), pre- and post-evaluation surveys (Figure 1B), and an aptitude assessment (Supplementary File S2). The objectives within the presentation, presented by these professors with extensive cancer research expertise, included:

**Figure 1:**
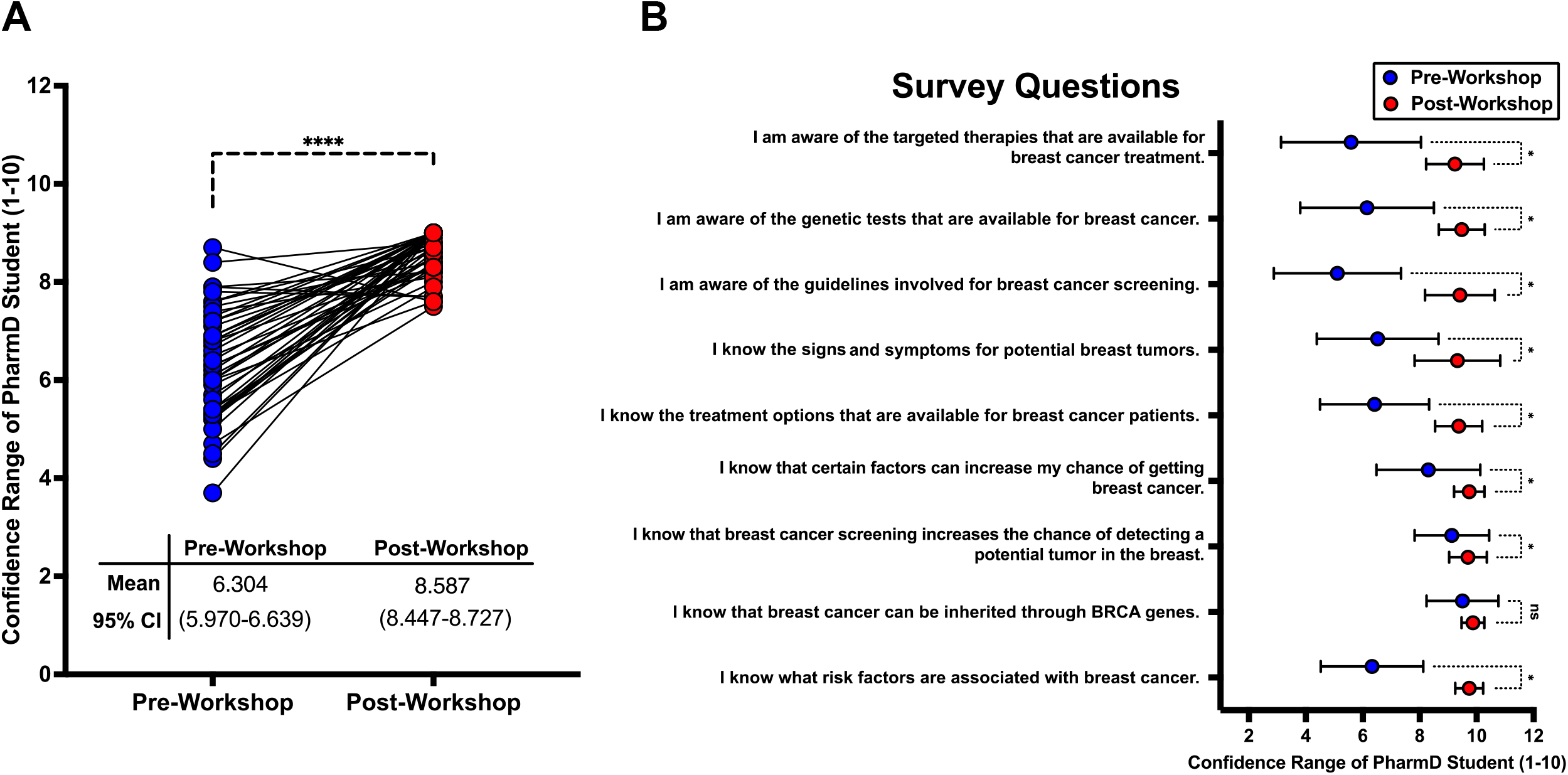
Impact of Train-the-Trainer Breast Cancer Educational Workshop on PharmD Students’ Confidence. **(A)** Comparison of PharmD students’ self-rated confidence levels (scale 1-10) before and after attending breast cancer educational workshop, with each line connecting to an individual student’s pre- and post-workshop confidence scores. The mean pre-workshop confidence was 6.304 (95% CI: 5.970-6.639) compared to post-workshop confidence of 8.587 (95% CI: 8.447-8.727; **** = p-value <0.0001, paired t-test). **(B)** Detailed breakdown of confidence ratings across nine specific breast cancer knowledge domains. Each question was rated on a scale of 1-10, with blue dots representing pre-workshop responses and red dots representing post-workshop responses. Error bars = 95% CI; * = p-value <0.05 (unpaired t-test) between pre- and post-workshop confidence changes for each topic, with “ns” indicating non-significant changes.

1. Describe the role of pharmacists in breast cancer education and prevention.
2. List modifiable and non-modifiable risk factors that can increase risk.
3. Describe various ways to reduce risk.
4. Explain the role genetics plays in the development of breast cancer.
5. Describe the different types/stages of breast cancer.
6. Explain various treatment options for patients who have been diagnosed.

The underlying purposes of these listed objectives were to increase overall awareness of breast cancer patient demographics, risk factors, staging, signs and symptoms, outcomes, and therapy options for this student cohort and subsequently for the community members who will receive these presentations.

The workshops were attended by 26 students in 2018 and 20 students in 2019 for a total of 46 participants. Students gave consent to participate in the study when signing up to attend the workshop. Pre- and post-workshop surveys were completed by each student, and results were collected to assess the overall efficacy of the workshop’s ability to provide then with confidence in understanding the workshop’s topics, such as breast cancer risk factors, signs and symptoms, screening, and treatment options. Furthermore, an aptitude assessment included multiple choice questions related to the material covered during the presentation was completed by each attendee immediately following the workshop, and an average score of 87.1 percent (95% CI 84.220-89.980) was obtained from our 2 cohorts. After completion of the workshop and final approval was provided by the presenters of the workshop, the PharmD student attendees were encouraged to present a similar structured presentation within the community at nearby community centers (for example, at churches, civic organizations, community health centers, *etc*.).

Statistics concerning the survey results were performed using GraphPad Prism 10 (GraphPad Prism Software, Boston MA). A paired t-test was performed on overall pre- and post-workshop survey scores; an unpaired t-test was performed for individual topics of the surveys; and a p-value threshold of 0.05 was used for determining statistical significance.

## RESULTS

To first address whether our train-the-trainer education workshop improved self-perceived confidence in our PharmD’s student cohort’s understanding in breast cancer risk factors, screening, signs and symptoms, and treatment options, we calculated the overall average confidence score for each administered survey to determine if post-workshop confidence scores significantly increased after completing the education workshop when compared to their corresponding pre-workshop confidence score. Based on our results, the de-identified PharmD student participants (n=46) exhibited a significant increase in confidence following the education training workshop, with their overall confidence range (on a scale of 1 to 10) improving from a pre-workshop average of 6.304 (95% CI 5.970-6.639) to a post-workshop average of 8.587 (95% CI 8.447-8.727) for all nine questions administered (paired t-test p-value <.0001) [Figure 1A]. This first result demonstrates that our train-the-trainer workshop was able to overall improve PharmD students’ confidence in understanding important facets of breast cancer risk and treatment options for the purposes of communicating this information to a local community.

To determine which topics of breast cancer education correlated with the best improvement in confidence for our PharmD student cohort, we performed a differential comparison for each survey question [Figure 1B]. Eight of the 9 survey questions corresponded with statistically significant improvement in confidence, with the most significant improvement being observed for the survey statement, “I am aware of the guidelines for breast cancer screening.” Specifically, participants’ confidence for this statement greatly increased from an average of 5.109 before the workshop to 9.413 afterward, reflecting a mean difference of 4.304. Conversely, the smallest change was noted for the statement, “I know that breast cancer can be inherited through BRCA genes.” The confidence in this statement was already high pre-workshop, at 9.500, and showed a slight increase to 9.870 post-workshop, with a mean difference of 0.3696. In fact, this survey statement was the only question that did not experience significant improvement in confidence post-workshop, compared to the other 8 questions in the surveys. Furthermore, the pre-workshop confidence in understanding BRCA gene inheritance appeared to be largely independent from the other genetics-related statement, “I am aware of genetic tests that are available for breast cancer.” The confidence in this statement improved from 6.152 to 9.478 in pre- and post-workshop surveys, respectively, reflecting a mean difference of 3.326. This disconnect between results was also seen in the statement, “I am aware of the target therapies that are available for breast cancer treatment,” with the 5.587 confidence pre-workshop improving to 9.239 post-workshop, with a mean difference of 3.652. These results are interesting to note, since the genomic/molecular status of breast cancers plays a large role in targeted therapy regimens. This disparity in responses to these seemingly related questions may suggest that while cultural awareness of BRCA gene inheritance is high, possibly due to media coverage regarding BRCA gene inheritance, it does not necessarily translate to an overall understanding of how genetic tests can impact breast cancer diagnosis and treatment regimens. These results showing increased improvement in comprehending genetics and targeted treatment topics in light of the potential disconnect in comprehending BRCA inheritance, along with the overall confidence improvements in comprehending topics regarding BRCA education among our PharmD students, demonstrate the utility this “train-the-trainer” program can provide to our PharmD students, who will then be able to similarly educate communities like those within rural Laurens, Greenwood, and Union Counties.

## DISCUSSION

In summary, our pilot program demonstrated the efficacy of a “train-the-trainer” model to enhance breast cancer education among PharmD students, ultimately aiming to enhance literacy and educational agency in rural communities and to contribute to reducing disparities in breast cancer outcomes within medically underserved communities. The “train-the-trainer” model’s workshop significantly increased student confidence and knowledge about breast cancer, as evidenced by substantial improvements in their post-versus pre-workshop survey scores. In fact, some of the largest improvements in confidence occurred regarding a topic centered around the themes of screening guidelines, application of genetic testing, and application of targeted therapies. This gain in knowledge is especially important for our PharmD students to promote education regarding genetic and targeted therapies in local rural communities, based on recent reports showing that rural practices have 80% lower odds of utilizing a molecular tumor board within a practice compared to those in urban settings, as well as less overall implementation of genomic tumor testing^20,21^. This approach highlights the potential for pharmacy professionals, who often serve as the most accessible healthcare professional for many patients, playing a pivotal role in bridging the gap in medical literacy and agency, especially in rural areas where access to quality healthcare information is limited.

The impact of this work extends beyond the immediate educational gains of the PharmD students. By empowering these future pharmacists with comprehensive knowledge and the confidence to educate others, we foster a sustainable model of community health education^22^. This initiative can lead to a culture that promotes earlier detection of breast cancer, more informed healthcare decisions, and better management of the disease, potentially decreasing mortality rates in underserved regions^6^. Additionally, the program exemplifies how targeted educational interventions can contribute to broader health equity, ensuring that critical health information reaches those who need it most. This study allows healthcare students who are located and working in rural areas to better understand a common disease and to have the ability to communicate these critical education points to a general population^22^. Exposure to resources and knowledge for diseases like breast cancer in rural communities through this program could also help catalyze more PharmD students to choose to work in disadvantaged areas^23^. The combined education they receive through schooling and the training program will not only equip them with the tools to be successful healthcare providers but also with the knowledge to better the geographical areas they choose to serve. While these pharmacists are encouraged to gain a deeper understanding of diseases, their role can expand by education and advocating for individual patients and their respective caregivers^24^.

With our promising results, this study has notable limitations. The sample size of participants is relatively small, which may limit the broader interpretation of the confidence measures reported by our PharmD student cohorts. Furthermore, the study relied on self-reported confidence measures and not less-subjective pre/post aptitude assessments, though an aptitude assessment was given. Future studies could include larger, more student diverse cohorts and incorporate long-term follow-up assessments with these students to determine the long-term effectiveness of this training as they gain experience presenting to communities. Looking ahead, future directions for this work could also focus on expanding the program to include other healthcare students and professionals through an interprofessional training environment (ex: physician assistants, nurses/nurse practitioners, occupational therapists, *etc*.), thereby broadening its reach and impact^25,26^. Additionally, this program for PharmD could also expand to other cancer types like cervical, colorectal, lung, and skin cancers that considerably impact rural communities^27,28^. Integrating objective assessments of knowledge retention and practical applications in community settings will provide even more comprehensive evaluation of the program’s effectiveness as seen in other health related train-the-trainer models^29^. Additionally, exploring approaches to address the identified gaps in baseline knowledge, such as enhanced focus on the scientific applications of genetic tests, will further strengthen the training. Ultimately, these efforts can help develop a robust framework for health education that empowers underserved communities, promotes health equity, and improves overall health outcomes of chronic diseases like breast cancer.

## Data Availability

All data produced in the present study are available upon reasonable request to the authors.

## ACKNOWLEDGEMENTS

We would like to thank Lisa Middleton, Director of Scientific Affairs at The Georgia Cancer Center in Augusta for her appreciated feedback and editing of this manuscript. AYS is supported by the National Institute of General Medical Sciences, the National Institutes of Health (SC INBRE P20GM103499).

## CONTRIBUTIONS

AYS and CLF conceived the project and designed workshop. MRG, ME, MMJ, AH, CA, MR, and HS coordinated participant recruitment and data collection. CNA and AYS analyzed the data. NAP, CLF, and AYS wrote the manuscript within input from all authors.

## ETHICAL DECLARATIONS

All human subjects-based work in this manuscript was performed in accordance with guidelines and regulations approved by the Institutional Review Board (IRB) of Presbyterian College.

## FIGURE LEGEND

**Supplementary File S1: PowerPoint Presentation of the PharmD Student BRCA Education Workshop**

**Supplementary File S2: Aptitude Assessment Questions Given to PharmD Student Post Workshop**

## Notes

### Competing Interest Statement

The authors have declared no competing interest.

### Author Declarations

All human subjects-based work in this manuscript was performed in accordance with guidelines and regulations approved by the Institutional Review Board of Presbyterian College.

