## Supplementary File S1 for "Leveraging Pharmacy Education through a Train-the-Trainer Model to Enhance Breast Cancer Literacy in Rural Communities"

### Breast Cancer & Genetics: Workshop Objectives

1) Describe the role of pharmacists in breast cancer education and prevention.
2) List modifiable and non-modifiable risk factors that can increase risk.
3) Describe various ways to reduce risk.
4) Explain the role genetics plays in the development of breast cancer.
5) Understand the different types/stages of breast cancer.
6) Explain various treatment options for patients who have been diagnosed.

### Goals of this Workshop

- Prepare students to educate the community on breast cancer risk, prevention, and treatment

- Improve knowledge of breast cancer in the community to promote healthier lifestyle choices, increase risk reduction, and encourage an active role in patients' own healthcare.

### Purpose of Workshop

- Continuation of research over the past 2 years to demonstrate that education in underserved communities about breast cancer can increase their knowledge, promote healthier choices, and empower their role in their personal healthcare

- Utilizing the “train-the-trainer” method allows pharmacy students to promote healthcare in the community, as well as keep the presentation information current.

### Why are we educating the community?

- **Goal:** Reduce the number of new cancer cases, as well as the illness, disability, and death caused by cancer.
- It's important to assess whether people understand and remember information received.
- Recommendations from health care providers are the most important reason patients point out for having screening tests.

*http://www.healthypeople.gov/2020/topics-objectives/topic/cancer?topicid=5*

### Female Breast Cancer
### Incidence Rates (per 100,000) by State

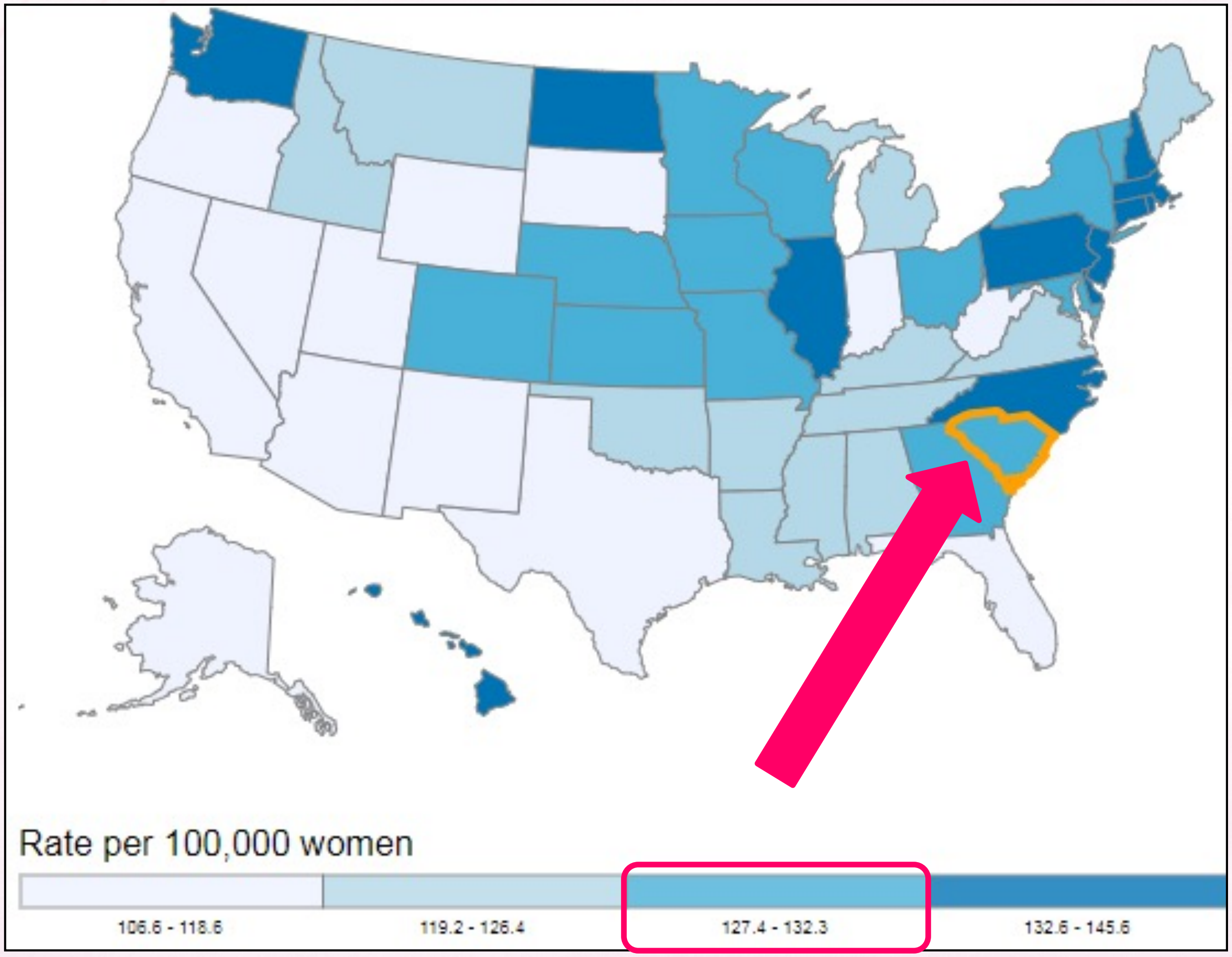


www.cdc.gov/uscs.

### Female Breast Cancer
### Death Rates (per 100,000) by State

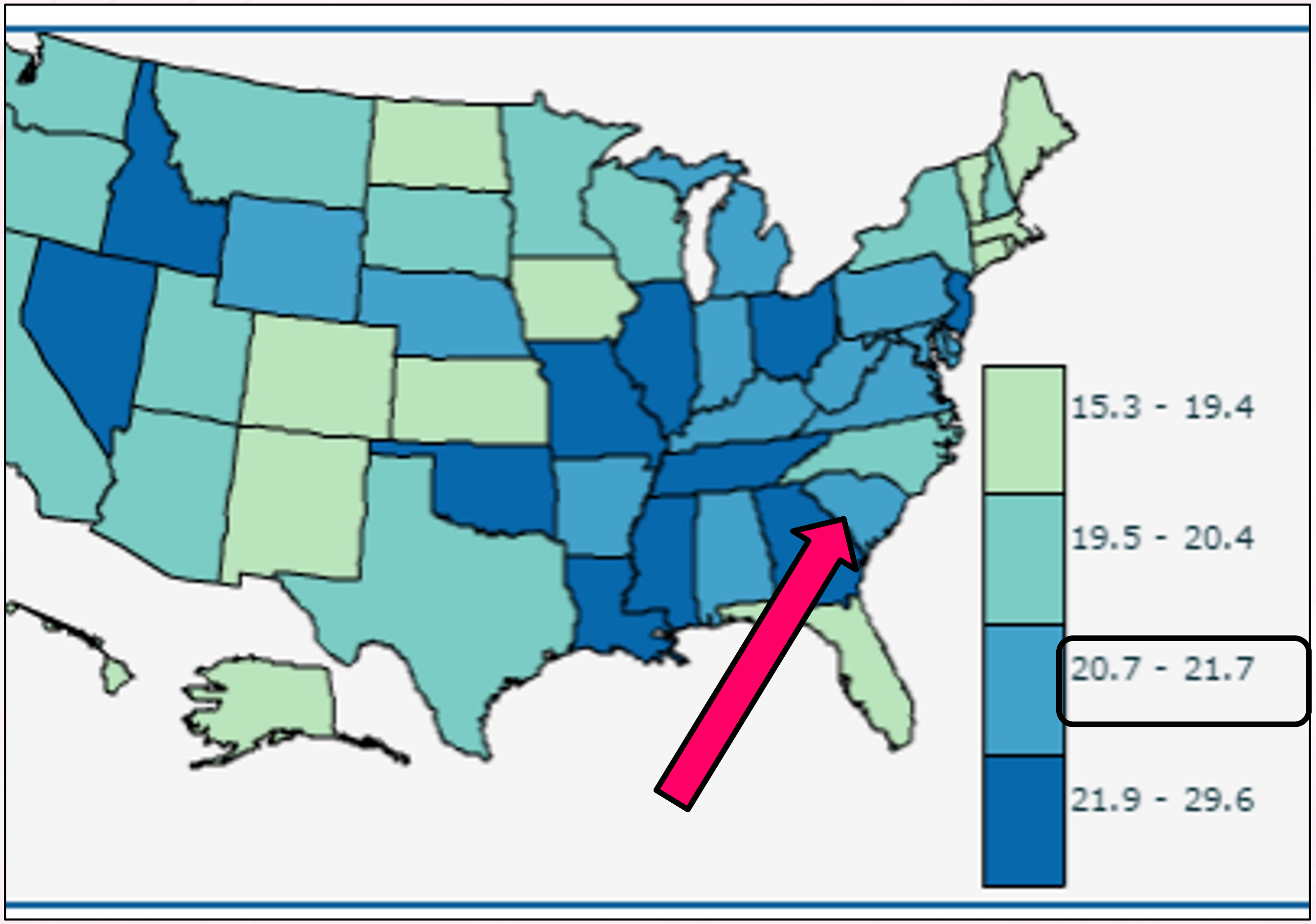


www.cdc.gov/uscs.

### What can pharmacists do to help?

- Most accessible providers for the population!
- Can educate:
  - Early screening recommendations
    - Self-breast examinations are KEY
  - Risks that increase chance of breast cancer
  - Medications
    - Treatment improvements
  - Signs & symptoms
  - Management
- Provide educational materials

***We want to give them the knowledge they need to work with their doctor on designing the best treatment for them or a loved one***

### Non-modifiable Risk Factors

1) **Gender:** female > male
2) **Age:** increased risk after age 55
3) **Race:** Caucasian rates > other races, but worse outcomes for Black/Hispanic
4) **Genetics:** increased risk if you have a 1st degree relative with breast or ovarian cancer; also BRCA1 and BRCA2 mutations increase risk
5) **PMH:** previous breast cancer increases risk of future breast cancer
6) **Reproductive History:** early menstruation and late menopause increases risk
7) **Breast tissue:** dense breast tissue increases risk

### Modifiable Risk Factors

1) **Physical activity:** exercising decreases risk
2) **Diet:** high fat and lack of fruits/veggies increases risk
3) **Weight:** obesity increases risk
4) **Alcohol:** more alcohol consumed=greater risk
5) **HRT:** combined hormone replacement therapy increases risk and advanced stage

### Socioeconomic Factors

- Breast cancer patients in lower-income areas have lower 5-year survival rates than those in higher-income areas at every stage of diagnosis

- Poverty, less education, and lack of health insurance are all associated with lower breast cancer survival

### Risk Assessment Tool

- Estimates a woman's risk of developing invasive breast cancer
- Explains common questions about breast cancer
- Designed by National Cancer Institute
  - http://www.cancer.gov/bcrisktool/
    - 5-year risk
    - Lifetime risk

### Reduce Risk of Development of Breast Cancer

- Lifestyle Changes
- Medications
- Early Screening

### Lifestyle Changes that can reduce risk

- Decreased alcohol consumption
- Lose weight if overweight or obese
- Increase physical activity
- Having a child before the age of 30 has been shown to decrease risk, as well as breastfeeding
- Avoid combined hormone replacement therapy if possible, as it can increase risk.

### Medications

**Approved for treatment and risk reduction:**

Tamoxifen (Soltamox)

Raloxifene (Evista)

- **Classification:** Selective Estrogen Receptor Modulators (SERMS)
- **MOA:** Block estrogen in some tissues and act like estrogen in others. In breast tissue, they block estrogen which fuels the growth of cancer cells, which is why they can be useful in lowering the risk.
- **Considerations:** Tamoxifen can be used by women whether or not they have gone through menopause, while raloxifene is only approved for use in postmenopausal women.
- **Duration of Therapy:** 5 years

### Medications

**Approved for treatment and used off-label for risk reduction**

Exemestane (Aromasin)

Letrozole (Femara)

Anastrazole (Arimidex)

- **Classification:** Aromatase Inhibitors
- **MOA:** Decrease estrogen levels by inhibiting aromatase in fat tissue from changing other hormones into estrogen. These drugs don’t stop the production of estrogen. They only lower estrogen levels in women whose ovaries aren’t making estrogen (i.e. menopause).
- **Considerations:** Studies are currently ongoing for the use of these drugs in risk reduction for breast cancer.

### Check Your Knowledge!

**Identify 5 modifiable risk factors associated with breast cancer:**

### Check Your Knowledge!

Identify 5 modifiable risk factors associated with breast cancer:

1. Physical inactivity
2. Poor diet
3. Overweight
4. Alcohol
5. HRT

### How is breast cancer detected?

- Breast Self Examination
- Clinical Breast Exam
- Mammography
- Ultrasonography
- Biopsy
- Fine needle aspiration
- Core biopsy
- Surgical biopsy
- Wire localization

### Signs & Symptoms

- Most important sign:
  - Lump/hard knot in breast or under-arm area (often painless)
- Swelling, warmth, redness of the breast
- Nipple change/change in breast size & shape
- Nipple discharge
- Dimpling or puckering of the skin
- Itchy/rash

### Signs & Symptoms

**The ACS recommends average risk patients start yearly mammograms starting at age 45, but all women should be aware of their normal anatomy and any changes should be discussed with their doctor.**

### Early Screening-Amy Robach

- In 2013, Good Morning America kicked off breast cancer awareness month special

- Amy Robach was asked to get mammogram on air to demystify the fear and nervousness for women that haven't been screened

http://abcnews.go.com/GMA/video/amy-robach-breast-cancer-diagnosis-mammogram-air-change-20848609

### Breast Cancer & Genetics

- The average woman in the US has about a 1 in 8 chance (12%) of developing breast cancer.
- About 5% to 10% of breast cancers are thought to be hereditary
- That chance increases to 80% in women who have an inherited abnormal BRCA1 or BRCA2 gene (or both).

### Cancer Genetics

- Cancer is a genetic disease which involves somatic alterations to the DNA of cells in a specific tissue (ex: breast) or certain cell types (ex: white blood cells)

- Tumor cells possess genes with somatic alterations that encode for mutated proteins involved in:
  - **Regulating or promoting cell cycle**
  - **DNA repair**

### Genetic markers

- **BR**east **CA**ncer genes: **BRCA 1** and **BRCA 2**
  - Human cancer suppressing genes
  - When mutated or altered, these genes cannot repair DNA damage, and further genetic alterations can occur that will lead to cancer.
- BRCA 1 **>** BRCA 2
  - Breast Cancer in Women
  - Ovarian Cancer in Women
- BRCA 2 **>** BRCA 1
  - Breast Cancer in Men

### Who should get tested for BRCA1 and BRCA2 mutations?

- The decision to be tested for mutations in BRCA1 or BRCA2 is best made in consultations with a genetic counselor and medical geneticist.

- The National Society of Genetic Counselors maintains a website that provides the names and locations of genetic counselors including those who specialize in cancer genetics. (http://www.nsgc.org/p/cm/ld/fid=164)

### Genetic testing

- Blood, swab, or saliva sample
- Recommended for:
  - ➢Newly diagnosed with breast / ovarian cancer
  - ➢STRONG family history of breast / ovarian cancer
- Costs vary widely
  - ➢Some insurance plans cover, including some Medicare plans
  - ➢Financial assistance programs available

### Genetic Testing Results

- Positive
  - Test detects a **known** harmful variation of BRCA1 or BRCA2 markers
    - Increased **risk** of developing **contralateral** breast cancer and ovarian cancer
- Negative
  - Test **does not** detect a known harmful variation of a BRCA marker
    - Risk of developing breast cancer is the same as that of the general population
- Ambiguous
  - Test shows a change in BRCA markers
    - The change has not been previously associated with breast cancer, so risk is unknown

### Check Your Knowledge!

Which genetic marker is associated with a higher incidence of breast and ovarian cancer in *women*? in *men*?

### Check Your Knowledge!

Which genetic marker is associated with a higher incidence of breast and ovarian cancer in *women?* in *men?*

- Women: BRCA 1 > BRCA 2
- Men: BRCA 2 > BRCA 1

### Types of breast cancer

#### Noninvasive

- in situ breast cancer
  - In localized part of breast
  - not spreading to tissues, lobules, ducts

#### Invasive

- infiltrating breast cancer
  - Spread to other parts of the body

### Staging of cancer

- **Staging of cancer is based on**:
  - Size of tumor
  - Number affected lymph nodes
  - Signs of cancer invading other organs

- **Types of Staging**
  - TNM
  - Stage 0-4

### TNM Staging

- **T** stands for the original (primary) **tumor**.
- **N** stands for **nodes**. It tells whether the cancer has spread to the nearby lymph nodes
- **M** stands for **metastasis**. It tells whether the cancer has spread to distant parts of the body

### T Staging

- **TX**- tumor can't be measured
- **T0**-no evidence of a primary tumor
- **Tis**-cells are only growing in the most superficial layer of tissue, without growing into deeper tissues.
  –*in situ* cancer or *pre-cancer*
- Numbers after the T (such as **T1, T2, T3, and T4**) might describe the tumor size and/or amount of spread into nearby structures. The higher the T number, the larger the tumor and/or the more it has grown into nearby tissues.

### N Staging

- **NX-**nearby lymph nodes cannot be evaluated.

- **N0**-nearby lymph nodes do not contain cancer.

- Numbers after the N (such as **N1, N2, and N3**) might describe the size, location, and/or the number of nearby lymph nodes affected by cancer. The higher the N number, the greater the cancer spread to nearby lymph nodes.

### M Staging

- **M0**-no distant cancer spread was found.
- **M1**-cancer has spread to distant organs or tissues (distant metastases were found).

### Stage 0

- Noninvasive (in situ)-no spreading
- Types of cancer diagnosed
  - Ductal carcinoma in situ
  - Lobular carcinoma in situ

### Stage I

- Earliest stage of invasive breast cancer
- Spread to surrounding breast tissue
- Stage I is divided into:
  - Stage IA: tumor is ≤2 cm; has not spread outside breasts
  - Stage IB: small clusters of cancer cells that are no more than 2 mm; found in lymph nodes

### Stage II

- Primary tumor is larger than 2 centimeters across **without** underarm lymph node involvement

- Primary tumor is larger than 2 but less than 5 centimeters across ***with*** lymph node involvement

### Stage III

- Primary tumor has increased in size (> 5 centimeters) or any size that has spread to the skin, chest wall, or internal mammary lymph nodes

- Primary tumor is any size that has spread to more than 10 axillary lymph nodes

### Stage IV

- The tumor has spread to places far away from the breast to an organ, such as bones, lungs, liver, brain, or distant lymph nodes

### Other Tumor Descriptors

- Grade of the tumor
  - Well differentiated vs. poorly differentiated
- Hormone receptor status of the tumor
  - Estrogen Receptor – ER
  - Progesterone Receptor – PR
- HER2 protein expression status
- Menopausal status

### 5-year Survival Rate

| Stage | 5-year survival Rate |
|---|---|
| 0 | 100% |
| I | 100% |
| II | 93% |
| III | 78% |
| IV | 22% |

***http://www.cancer.org/cancer/breastcancer/detailedguide/breast-cancer-survival-by-stage***

### Check Your Knowledge!

**TRUE or FALSE?**

Determination of the tumor's expression of estrogen receptor (ER), progesterone receptor (PR), and HER2 protein is important when determining appropriate treatment.

### Check Your Knowledge!

**TRUE or FALSE?**

Determination of the tumor's expression of estrogen receptor (ER), progesterone receptor (PR), and HER2 protein is important when determining appropriate treatment.

**TRUE**

### Treating Breast Cancer: Combination of Local and Systemic Interventions

#### Local Interventions

- **Surgery**
  - partial or total mastectomy/BCT
- **Radiation**
  - may be used in large tumors or axillary lymph node involvement

#### Systemic/Adjuvant

- **Traditional Chemotherapy**
  - anthracyclines, taxanes, antimetabolites
- **Endocrine Therapy**
  - aromatase inhibitors, SERMs, HRT
- **Targeted & Biologic Therapy**
  - anti-HER2 monoclonal antibodies/tyrosine kinase inhibitors

### Treating Noninvasive Breast Cancer Stages 0-IA

**OFTEN TREATED WITH SURGERGY OR RADIATION, BUT NOT CHEMOTHERAPY**

### Combined Treatment Approach for Invasive Breast Cancer Stages IB-IV

#### Adjuvant Approach

1. **Local Surgery +/- Radiation**
2. **Systemic Adjuvant Therapy**
   – chemotherapy or targeted therapy used after surgery and/or radiation to reduce the risk of recurrence

#### Neoadjuvant Approach

1. **Systemic Adjuvant Therapy**
   – chemotherapy or targeted therapy used before local interventions to improve success of tumor resection
2. **Local Surgery +/- Radiation**

***Treatment approach is dependent upon likelihood of recurrence, surgical margins, tumor size, axillary lymph node involvement, and HER2 status of the tumor.***

### Traditional Chemotherapy

- Anthracyclines-damage the genetic material of cancer cells → cancer cells die.
  - **Adriamycin, Ellence, daunorubicin**

- Taxanes-interfere with the way cancer cells divide
  - **Taxol, Taxotere, Abraxane**

- Antimetabolites- kill cancer cells by acting as false building blocks in a cancer cell's genes → cancer cells die as they get ready to divide
  - **Gemzar, methotrexate, Xeloda, Adracil**

### Targeted Therapy

- Somatic alterations in tumor cells create different proteins (receptors) than normal cells
- Drugs (messengers) can be developed that target the altered or amplified receptors
  - More effective and individualized treatment
  - Fewer side effects

### Approved Targeted Therapy

- **HER2-positive** breast cancer overexpresses the protein(receptor) human epidermal growth factor receptor 2 due to a gene mutation ("cell flags")
- This receptor results in longer life and growth of the cell
- Hospitals can identify in the clinical labs using special test kits

### Approved Targeted Therapy

#### Injectable Monoclonal Antibody Inhibitors

- **Trastuzumab (Herceptin)**
  - Targets the HER2 protein receptor
  - Used in combination with chemotherapy

- **Pertuzumab (Perjeta)**
  - Targets the HER2 from dimerizing  and activating the receptor
  - Used in combination with chemotherapy & Trastuzumab

- **Ado-trastuzumab emtansine (Kadcyla)**
  - Humanized antibody that targets the HER-2 protein receptor
  - Used to treat HER2-positive cancer patients who have metastatic or a cancer that has recurred

### Approved Targeted Therapy

#### Small Kinase Inhibitor

- **Lapatinib (Tykerb)**
  - Binds to the tyrosine kinase of the EGF receptors (EGFR & HER2).
  - Used in combination with chemotherapy
    - Examples: **Capecitabine (Xeloda)** or **letrozole (Femara)**
  - Treat HER2 positive patients who did not respond to the first line of treatment.

### Approved Targeted Therapy

- **Everolimus (Afinitor)**
  - ER–positive, HER2/neu–negative breast cancers failing to respond to letrozole
  - Several proteins have been mutated in the AKT1 pathway of breast tumors.
  - Targets the mTOR protein by inhibiting the protein which promotes the growth of the tumor and cell division.

### Online resources for more information

- National Cancer Institute (NCI)
  - www.cancer.gov
  - Cancer Topics
  - Clinical Trials
  - Cancer Statistics

- Breast Cancer Organization
  - http://www.breastcancer.org
  - Symptoms and Diagnosis
  - Treatment & Side Effects

- Susan G. Komen
  - http://ww5.komen.org/BreastCancer/AboutBreastCancer.html
  - Facts & Information
  - Get Involved

- Greenwood Genetic Center
  - http://www.ggc.org/
  - Testing and counseling

### What Now?

Come to Workshop

Take Follow-up Quiz in Moodle

Present Community Presentation to Dr. Farrell, Melanie Routhieaux, or Morgan Enlow as checkoff

Go out into the community and give the presentation
