## Supplementary File S2 for "Leveraging Pharmacy Education through a Train-the-Trainer Model to Enhance Breast Cancer Literacy in Rural Communities"

### APTITUDE ASSESSMENT FOR BRCA EDUCATION WORKSHOP

1. South Carolina has one of the highest cases of breast cancer in the country. T/F
   A. True
   B. False

2. Which race/ethnic group has a higher rate of breast cancer diagnosis?
   A. Caucasians
   B. African Americans
   C. Hispanics
   D. Asians

3. Which of the following socioeconomic factors is associated with lower breast cancer survival (5 year survival)?
   A. Poverty
   B. Less education
   C. Lack of health insurance
   D. All of the above

4. At what age should one start beginning annual mammograms?
   A. 55
   B. 45
   C. 25
   D. 60

5. Which of following is associated more with the variants in the BRCA2 gene over the BRCA1 gene?
   A. Breast Cancer in Women
   B. Breast Cancer in Men
   C. Ovarian Cancer in Women
   D. All of the above

6. What are some lifestyle changes that can decrease the risk for breast cancer?
   A. Decreased alcohol intake
   B. Decreased weight
   C. Decreased physical activity
   D. A and B

7. What medications are approved to reduce the risk for breast cancer for certain patients groups?
   A. Tamoxifen
   B. Elavil
   C. Raloxifene
   D. A and C

8. Raloxifene is approved for pre-menopausal women? T/F
   A. True
   B. False

9. Staging of cancer is based upon which of the following:
   A. Size of tumor
   B. Number of affected lymph nodes
   C. Signs of cancer invading other organs
   D. All of the above

10. The M for TNM staging stands for:
   A. Meta-analysis
   B. Metastatic
   C. Muscle cancer
   D. Melanoma

11. What is the treatment needed for stages 0 and 1?
   A. Surgery
   B. Radiation
   C. Chemotherapy
   D. A and B
   E. A, B, and C

12. What is an example of targeted therapy?
   A. Trastuzumab (Herceptin)
   B. Lapatinib (Tykerb)
   C. Everolimus (Afinitor)
   D. A and B
   E. A, B, and C
